# Prevalence, treatment rates and control of dyslipidemia among diabetes patients in northern Nigeria: A cross-sectional, multicenter study

**DOI:** 10.64898/2026.03.19.26348864

**Authors:** Nura Hamidu Alkali, Andrew Enemako Uloko, Godwin O. Osaigbovo, Adamu Girei Bakari, Mohammed Raji Bello, Maria Ahuoiza Garba, Garba Musa Fika, Abubakar Sahabi Muhammad, Muhammad Arabi Saad, Zira Ghyi Vandi, Umar Faruk Abdullahi, Ayuba Mugana, Ijuptil Chiroma, Ibrahim Abdullahi Haladu, Lumsami Shadrach, Usman Adamu Nuhu, Godiya Ishaya Dare

**Author notes:** Corresponding author: (NHA).

## Abstract

**Objectives:** Dyslipidemia is prevalent among Nigerians living with diabetes, but the rate and extent of treatment have not been well-studied. The objective of this study was to determine the prevalence, treatment rates and control of dyslipidemia among diabetes patients in northern Nigeria.

**Methods:** We conducted a multicenter, cross-sectional study of diabetes clinic patients. We noted cardiovascular risk factors, lipid-lowering treatments and examination findings, including body mass index, blood pressure, glycated hemoglobin, lipid profile, glomerular filtration rate and urinalysis. Outcome measures were the rate of dyslipidemia, the proportion of patients treated for dyslipidemia, and the proportion of patients with low density lipoprotein cholesterol goal and target for primary prevention of cardiovascular disease.

**Results:** The study enrolled 403 participants (58.8% females), of whom 59.6% had dyslipidemia. Female gender and proteinuria were independently associated with dyslipidemia, with odds ratios of 1.74 and 2.26, respectively. Other cardiovascular risk factors of participants were hypertension (56.8%), obesity (52.6%), chronic kidney disease (36.5%), atrial fibrillation (7.9%), heart failure (5.0%), cigarette smoking (4.7%), excess alcohol use (2.0%), and previous cardiovascular disease (14.4%). In those with dyslipidemia, 51.3% took lipid-lowering treatments comprising statins (49.6%), clofibrate (1.7%) and statins combined with clofibrate (1.2%). None took other lipid-lowering treatments beside dietary control, probably due to high costs compared to statins. Only 17.1% of all participants attained the target for primary prevention of cardiovascular disease in people with diabetes.

**Conclusion:** Most patients had dyslipidemia, which was more prevalent in females. Only a sixth of all patients had attained the treatment target. Treatment for dyslipidemia was limited to statins and fibrates, contrary to guideline recommendations for the use of ezetimibe, bempedoic acid, icosapent ethyl, or PCSK9 inhibitors for those who failed intensive statin therapy. There is a need for physician adherence to practice guidelines for the treatment of dyslipidemia, and improved access to treatment in northern Nigeria.

## Introduction

Diabetes mellitus (DM) is a chronic metabolic disease and a major cause of death and disability worldwide [1]. Up to 6.1% of the global adult population had DM in 2021, which was expected to rise to 10% or more in developing countries by 2050 [2]. DM prevalence in children is also rising worldwide. The Global Burden of Disease study has found that the prevalence of childhood DM rose by 39.4% between 1990 and 2019, with the highest rise seen in low-income countries [3].

DM is associated with high risks of cardiovascular diseases (CVD), including stroke, myocardial infarction, peripheral arterial disease (PAD), heart failure and pulmonary thrombo-embolism (PTE).^1^ Indeed, DM contributed 20% to the population attributable risk of stroke in Nigeria, while a UK study found CVD events and mortality were two to three-fold higher among DM patients than the general population [4,5]. In other studies, DM patients with no history of CVD had a similar risk of myocardial infarction as non-DM subjects with a history of CVD, suggesting that CVD risk due to DM is equivalent to a previous CVD [6]. The high CVD risks associated with DM result from premature atherosclerosis complicating the chronic effects of hyperglycemia, dyslipidemia, hypertension, obesity, and other metabolic derangements seen in DM patients [7]. For instance, in the Action for Health in Diabetes (Look AHEAD) study, DM patients with dyslipidemia had 1.3 and 1.5 times additional risks of stroke and coronary heart disease (CHD) compared to those with DM alone [8]. Other effects of dyslipidemia in people with DM are retinopathy, macular edema, erectile dysfunction, diabetic foot syndrome and fetal anomalies during pregnancy [9–11].

Dyslipidemia is defined as high levels of serum low density lipoprotein cholesterol (LDLC), total cholesterol (TC) and/ or triglycerides (TG), or low levels of high density lipoprotein cholesterol (HDLC) [12]. The major determinants of human lipoprotein levels are dietary intake, obesity, insulin resistance and genetic factors influencing lipid absorption, transport and metabolism [12]. Among genetic factors causing high LDLC are defects of the enzyme lipoprotein lipase that degrades LDL-rich TGs and very low density lipoproteins (VLDL) within chylomicrons [13,14]. Regardless of underlying cause, excess LDLC activates monocytes and macrophages to release pro-inflammatory cytokines, including vascular cell adhesion molecule-1 and interleukin-1, that further stimulate macrophages to engulf LDLC, transform into foam cells and form atherogenic plaques in the tunica media [7,13]. Platelet aggregation at the sites of atherogenic plaques subsequently causes vascular thrombosis. Raised serum levels of apolipoproteins A and B also promote thrombogenesis. While apolipoprotein A promotes the growth of atherogenic plaques, apolipoprotein B competes with plasminogen and tissue plasminogen activator for binding to fibrin, thereby inhibiting plasmin-mediated fibrinolysis [14,15].

The prevalence of dyslipidemia in people with DM ranges from 43.5% among type 1 DM patients in Spain to 95% among type 2 DM patients in Jordan [16,17]. The prevalence rate is as high as 72.6% in parts of Nigeria, although the average rate across sub-Saharan Africa is 52.7% [18,19]. Thus, cardiovascular risk reduction in DM patients with dyslipidemia is a major goal of DM care worldwide [18–20]. Current practice guidelines recommend the use of statins as first-line treatment for dyslipidemia. The Cholesterol Treatment Trialists’ Study has shown that use of statin to lower raised LDLC caused proportionate reductions in coronary events and stroke, while the ODYSSEY trial showed that add-on therapy with a PCSK9 inhibitor caused fewer rates of ischemic stroke, transient ischemic attack (TIA), CHD and fatal myocardial infarction [20–22]. Other studies support the use of niacin, fibrates, ezetimibe and bempedoic acid as alternate or add-on therapies for those who failed intensive treatment with statins or experience intolerable side effects.^23^ Accordingly, clinical practice guidelines recommend statins and other lipid-lowering treatments (LLTs) to prevent CVD in DM patients with dyslipidemia [23–25]. Yet, evidence of this practice is scarce in Nigeria, where several studies have assessed the prevalence and patterns of dyslipidemia in DM patients, but not the rates or outcomes of statin therapy [19,26–29]. A study from Nigeria also found that only 19% of stroke patients with dyslipidemia had taken LLTs, suggesting poor care of dyslipidemia among other Nigerians at high risk of CVD [30].

In this study, we aimed to determine the prevalence and treatment rates of dyslipidemia among DM patients in northern Nigeria, and the proportion of patients that attained the LDLC goal and target recommended for the primary prevention of CVD. Findings from this study could improve the care of dyslipidemia among Nigerians living with DM.

## Materials and Methods

### Study design and setting

We conducted a cross-sectional, observational study of CVD risks and metabolic outcomes among patients receiving care at the DM clinics of Abubakar Tafawa Balewa University Teaching Hospital Bauchi (ATBUTH), Ahmadu Bello University Teaching Hospital Zaria (ABUTH), Aminu Kano Teaching Hospital Kano (AKTH), Modibbo Adama University Teaching Hospital Yola (MAUTH), and the University of Maiduguri Teaching Hospital (UMTH), Maiduguri, Nigeria. The study was conducted from 8^th^ February to 15^th^ December, 2023. Findings on CVD risks, exercise practices and metabolic outcomes were published previously [31].

The present study utilized data on the prevalence and treatment rates of dyslipidemia, and the proportion of patients, who at enrolment, had attained the LDLC goal and target recommended for the primary prevention of CVD during previous care of DM. The study sites were located in Bauchi, Kaduna, Kano, Adamawa and Borno states of Nigeria, a country of 923,768 square kilometers comprising 36 states and a Federal Capital Territory. The DM clinics at study sites operated once or twice weekly, and were all staffed by physician diabetologists, physiotherapists, nutritionists, DM care nurses and counsellors. Each clinic had point-of-care testing for glycated hemoglobin (HbA1c), fasting blood glucose (FBG) and urinalysis.

### Study population and sampling

Participants were recruited consecutively after signing a written, informed consent. Parents or guardians signed consent and filled study questionnaires for those patients aged below 18 years. The sample size was calculated using Cochran’s formula for unknown proportions: *n* = Z^2^ x p (1-p)/e^2^, where *n* is the minimum required sample size, *Z* is the standard normal (1.96) corresponding to a 95% confidence interval (CI), *p* is the unknown proportion of DM patients with dyslipidemia in the study area (0.5), and *e* is the margin of error, placed at 5%.^32^ Thus, the minimum required sample size was *n* = (1.96)^2^ x 0.5(1 – 0.5)/0.05^2^ = 385. We anticipated a 4% attrition rate and added 16 participants, which raised the final sample size to 401.

### Inclusion and exclusion criteria

Patients were included in the study if they consented to participate, had attended the DM clinic at least once, and attended at least two follow-up visits during the study period. Those who missed a second follow-up visit during the study period or failed to perform lipid profile tests were excluded from the study.

### Study instruments and variables

Data were collected on a standard questionnaire and pro-forma form that noted participants’ sociodemographic and past medical history, as well as duration of DM diagnosis, type of DM, complications of DM, history of dyslipidemia, hypertension, heart failure, atrial fibrillation, chronic kidney disease (CKD), angina, myocardial infarction (MI), stroke, PAD, PTE, illicit drug use, cigarette smoking and excess use of alcohol. Other items were the treatment history, including the use of insulin, non-insulin injectables, oral glucose-lowering drugs (GLD), antihypertensive drugs, statins, fibrates, niacin, ezetimibe, bempedoic acid, inclisiran, icosapent ethyl, PCSK9 inhibitors and other LLTs. Examination and laboratory findings included weight and height measurements, body mass index, waist circumference, blood pressure (BP), serum FBG, HbA1c, TC, LDLC, HDLC, TG, electrolytes, urea, creatinine and urinalysis. The glomerular filtration rate (GFR) was calculated from participant’s creatinine values, age and sex using an online eGFR calculator of the U.S. National Kidney Foundation (https://www.kidney.org/professionals/gfr_calculator). An eGFR ≤ 59 ml/ minute/ 1.73 M^2^ body surface area, constituted CKD and also a risk factor for CVD [15].

### Data collection procedures

Weight and height were measured with the participant standing on a stadiometer, wearing light clothing and no shoes. The BMI was calculated from the body weight in kilograms divided by the square of the height in meters. Waist circumference was measured with a non-stretching tailor’s tape placed midway between the lower rib margins and the anterior superior iliac spines. Obesity was defined as a BMI of 30 kg/M^2^ or more, or waist circumference reaching 102 cm in males or 88 cm in females. The BP was the average of three readings taken with a mercury sphygmomanometer applied to the arm. Blood and urine specimens were sampled at the DM clinic after an overnight fast and tested for FBG, HbA1c and urinalysis, or sent to the main hospital laboratory for tests on lipid profile, electrolytes, urea and creatinine.

### Operational terms and definitions

Type 1 and type 2 DM were defined according to the methods of Rawshani et al [33]. Type 1 DM was treatment with insulin and DM diagnosis at age < 30 years, and type 2 DM was treatment with diet, with or without oral GLD, or treatment with insulin with or without oral GLD in patients who were 40 years or older at DM diagnosis. HbA1c values ≤ 7.0% was defined as good glycemic control and higher values as poor control. Excess use of alcohol was defined as alcohol intake more than 56 grams daily or 196 grams weekly in men, or 26 grams daily/ 98 grams weekly in women [34]. Dyslipidemia was defined as serum TC > 6.2 mmol/L, LDLC > 4.1 mmol/L, TG ≥ 2.3 mmol/L or HDLC < 1.0 mmol/L in males or 1.3 mmol/L in females, according to the National Cholesterol Education Program-Adult Treatment Panel III guidelines [35]. Normal values excluded dyslipidemia regardless of history.

### Outcome measures

The main outcome measures were the: 1. Rates and patterns of dyslipidemia, 2. Proportion of patients treated for dyslipidemia, and 3. Proportion of patients, who at enrolment, had attained the LDLC goal and target of < 1.8 mmol/L for primary prevention of CVD in people with DM as recommended by the European Society of Cardiology/ European Atherosclerosis Society [15].

### Statistical analysis

Data were analyzed with the Statistical Package for the Social Sciences (SPSS) version 25 program (IBM Corporation, New York, USA). Categorical variables were analyzed with the Pearson chi-square test or Fisher’s exact test, while means of continuous variables were analyzed with the Student’s *t* test. Binomial logistic regression was used to analyze independent associations between outcome measures and potential participant characteristics implicated in previous studies. P values less than 0.05 were considered statistically significant. A four-set Venn diagram was used to show intersections of low HDLC, high TC, high LDLC and high TG counts defining dyslipidemia [36]. Results were presented as proportions, means, medians, odd ratios (OR), standard deviations (SD) and 95% confidence intervals (CIs) in tables and text. The methods and results were reported according to the guideline protocol of “Strengthening the Reporting of Observational Studies in Epidemiology” (STROBE) [37].

## Ethical considerations

The study was approved by the Health Research Ethics Committees of ATBUTH Bauchi (Ref. 005/2023, dated January 11, 2023), ABUTH Zaria (Ref. ABUTHZ/HREC/F41/2023, dated January 12, 2023), UMTH Maiduguri (Ref. OHRP-IRB00013572 UMTH/REC/23/ 1110, dated January 31, 2023), MAUTH Yola (Ref. HREC/23/239, dated February 3, 2023), and AKTH Kano (NHREC/28/01/2020/AKTH/EC/3532, dated March 14, 2023). The study protocol has conformed to the Helsinki Declaration.

## Results

### Demographic and clinical characteristics

We enrolled 403 patients (58.8% females) out of 426 screened for the study. Thus, the response rate was 94.6% (Fig 1).

**Fig 1:**
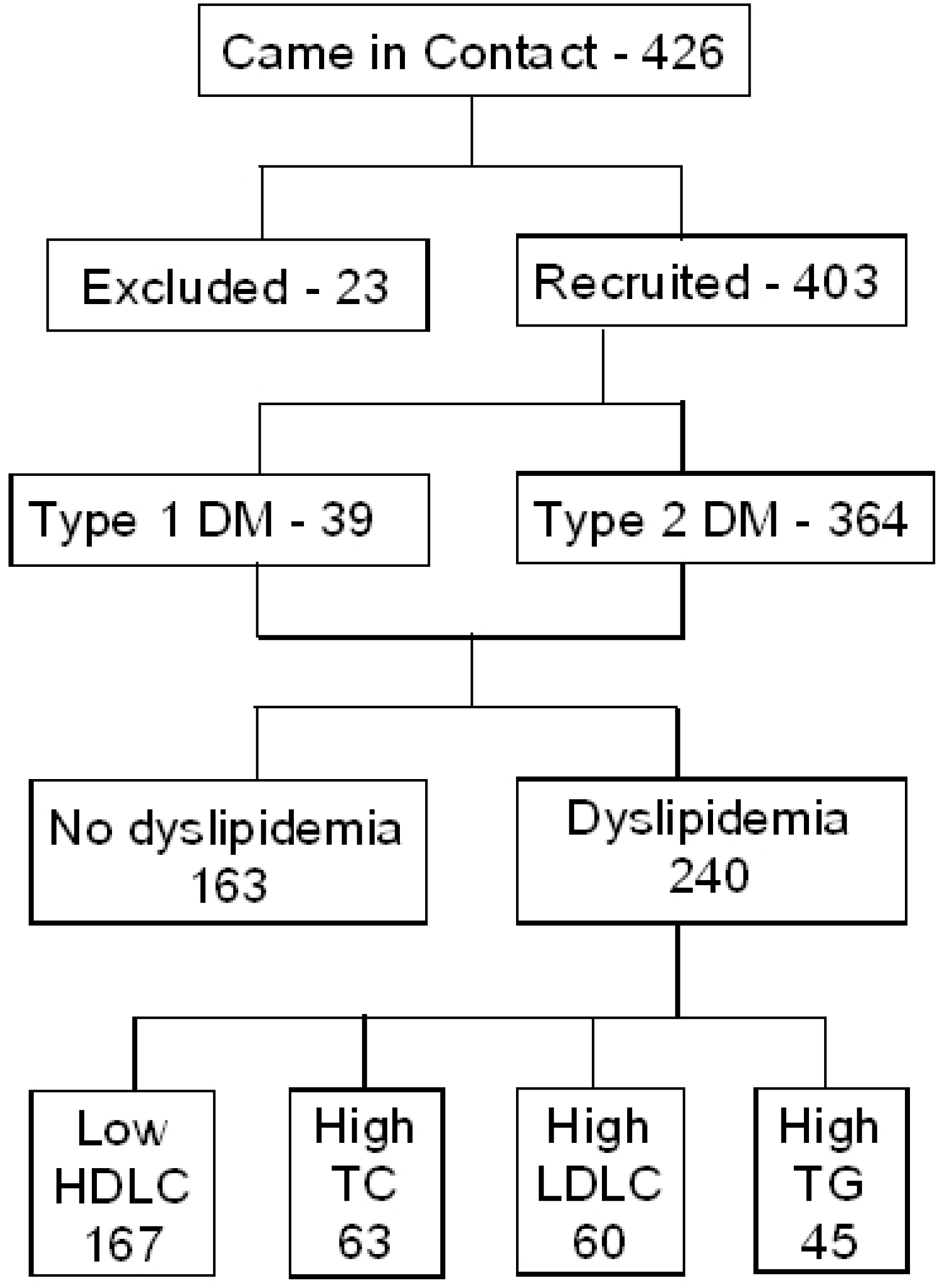
Flowchart of study participants. DM, Diabetes mellitus; HDLC, High density lipoprotein cholesterol; LDLC, Low density lipoprotein cholesterol; TC, Total cholesterol; TG, Triglycerides

Mean age of participants (±SD) was 53.2±12.7 years, with males being significantly older than females (54.9±11.7 years vs. 52.1±13.3 years, respectively; 95% CI: 0.35–5.39, *P* = 0.026) and two-thirds participants being urban residents. Other demographic characteristics are shown on Table 1. Those excluded from the study were similar to participants in age (mean age ± SD, 47.8±16.7 vs. 53.2±12.7 years, respectively; 95% CI: –0.05–10.89; *P* = 0.052), gender ratio (females, 56.5% vs. 58.8%, respectively; χ^2^ = 0.047, *P* = 0.83) and place of residence (urban area, 78.3% vs. 70.0%, respectively; χ^2^ = 0.72, *P* = 0.49). Beside DM and dyslipidemia, CVD risk factors also included hypertension (56.8%), obesity (52.6%), CKD (36.5%), atrial fibrillation (7.9%), heart failure (5.0%), cigarette smoking (4.7%), excess alcohol use (2.0%), and a past history of CVD (14.4%). Other clinical and laboratory characteristics of participants are shown on Table 2.

**Table 1:** Comparison of participant demographic characteristics stratified by gender.

| **Characteristic** | **Total**<br>**(N = 403)** | **Male**<br>**(n = 166)** | **Female**<br>**(n = 237)** | **$\chi^2$**<br>**Statistic** | **P-Value** |
| --- | --- | --- | --- | --- | --- |
| Study site, *n* (%) |  |  |  |  |  |
| Bauchi | 64 (15.9) | 30 (18.1) | 34 (14.3) | 3.06 | 0.55 |
| Kano | 117 (29.0) | 42 (25.3) | 75 (31.6) |  |  |
| Maiduguri | 77 (19.1) | 35 (21.1) | 42 (17.7) |  |  |
| Yola | 75 (18.6) | 29 (17.5) | 46 (19.4) |  |  |
| Zaria | 70 (17.4) | 30 (18.1) | 40 (16.9) |  |  |
| Place of residence, *n* (%) |  |  |  |  |  |
| Urban area | 282 (70.0) | 110 (66.3) | 172 (72.6) | 1.85 | 0.186 |
| Rural area | 121 (30.0) | 56 (33.7) | 65 (27.4) |  |  |
| Educational qualification, *n* (%) |  |  |  |  |  |
| Primary school certificate | 45 (11.2) | 8 (4.8) | 37 (15.6) | 30.63 | < 0.001* |
| Secondary school certificate | 93 (23.1) | 43 (25.9) | 50 (21.1) |  |  |
| University degree/diploma | 154 (38.2) | 76 (45.8) | 78 (32.9) |  |  |
| Postgraduate qualification | 37 (9.2) | 22 (13.25) | 15 (6.3) |  |  |
| No formal education | 74 (18.4) | 17 (10.2) | 57 (24.1) |  |  |
| Occupation, *n* (%) |  |  |  |  |  |
| Student/ Youth Service Corps | 13 (3.2) | 2 (1.2) | 11 (4.6) | 36.94 | < 0.001* |
| Farmer/ trader/ artisan | 64 (15.9) | 26 (15.7) | 38 (16.0) |  |  |
| Civil servant/ public servant | 111 (27.5) | 55 (33.1) | 56 (23.6) |  |  |
| Private company employee | 12 (3.0) | 3 (1.8) | 9 (3.8) |  |  |
| Police, military, paramilitary | 13 (3.2) | 12 (7.2) | 1 (0.4) |  |  |
| Business person/contractor | 56 (13.9) | 31 (18.7) | 25 (10.5) |  |  |
| Retiree/unemployed/FTHW | 134 (33.25) | 37 (22.3) | 97 (40.9) |  |  |
| Marital status, *n* (%) |  |  |  |  |  |
| Single | 22 (5.5) | 8 (4.8) | 13 (5.5) | 33.05 | < 0.001* |
| Married | 320 (79.4) | 153 (92.2) | 168 (70.9) |  |  |
| Divorced/Widowed | 61 (15.1) | 5 (3.0) | 56 (23.6) |  |  |

\* Significant p-value; FTHW, Full-time housewife

**Table 2:** Cardiovascular risk factors among participants compared by χ^2^ and Student *t* tests.

| **A. Categorical variables, n (%)** | **Total (N = 403)** | **Male (n = 166)** | **Female (n = 237)** | **$\chi^2$ Statistic** | **P-Value** |
| --- | --- | --- | --- | --- | --- |
| Type of DM |  |  |  |  |  |
| Type 1 | 39 (9.7) | 11 (6.6) | 28 (11.8) | 3.0 | 0.09 |
| Type 2 | 364 (90.3) | 155 (93.4) | 209 (88.2) |  |  |
| Old age ($\geq 65$ years) | 79 (19.6) | 32 (19.3) | 47 (19.8) | 0.02 | 1.0 |
| Obesity | 209/397 (52.6) | 48/162 (29.6) | 161/235 (68.5) | 14.85 | < 0.001* |
| Dyslipidemia | 240 (59.6) | 84 (50.6) | 156 (65.8) | 9.389 | 0.003* |
| Hypertension | 229 (56.8) | 89 (53.6) | 140 (59.1) | 1.18 | 0.307 |
| Atrial fibrillation | 32 (7.9) | 15 (9.0) | 17 (7.2) | 0.464 | 0.575 |
| Heart failure | 20 (5.0) | 9 (5.4) | 11 (4.6) | 0.126 | 0.817 |
| Cigarette smoking | 19 (4.7) | 19 (11.4) | 0 (0.0) | 28.5 | < 0.001* |
| Excess use of alcohol† | 8 (2.0) | 7 (4.2) | 1 (0.4) | 7.23 | 0.01* |
| Past history of CVD | 58 (14.4) | 22 (13.2) | 36 (15.2) | 0.3 | 0.67 |
| Chronic kidney disease | 142/389 (36.5) | 52/161 (32.3) | 90/228 (39.5) | 2.1 | 0.165 |
| Proteinuria | 160/402 (34.8) | 67 (40.4) | 93/236 (39.4) | 0.037 | 0.918 |
| **B. Continuous variables, Mean (SD)** | **Total (N = 403)** | **Male (n = 166)** | **Female (n = 237)** | **95% CI** | **P Value** |
| Age, years | 53.2 (12.7) | 54.9 (11.7) | 52.1 (13.3) | 0.35 – 5.39 | 0.026* |
| Duration of DM, months | 100.4 (85.3) | 104.0 (88.2) | 97.8 (83.4) | - 4.8 – 23.1 | 0.48 |
| Body mass index, kg/M<sup>2</sup> | 28.5 (5.8) | 26.4 (4.8) | 29.9 (6.1) | 2.3 – 4.7 | < 0.001* |
| WC, centimeter | N/A | 94.5 (13.1) | 95.7 (15.4) | N/A | N/A |
| Systolic BP, mm/Hg | 131.7 (18.9) | 132.6 (18.5) | 131.2 (19.2) | -2.82 – 5.71 | 0.51 |
| FBG, mmol/L | 8.8 (4.1) | 8.5 (4.0) | 9.0 (4.1) | -1.31 – 0.3 | 0.22 |
| HbA1c, % | 8.7 (2.1) | 8.6 (2.1) | 8.8 (2.2) | -0.69 – 0.2 | 0.27 |
| Serum creatinine, μmol/L | 117.3 (81.3) | 128.7 (96.5) | 109.2 (68.4) | 3.1 – 36.0 | 0.02* |
| GFR, ml/minute/1.73M<sup>2</sup> | 72.2 (30.4) | 73.4 (29.1) | 71.4 (31.3) | -4.14 – 8.16 | 0.52 |
| Serum lipid profile, mmol/L |  |  |  |  |  |
| Total cholesterol | 5.0 (1.4) | 4.9 (1.4) | 5.1 (1.3) | -0.42 – 0.13 | 0.3 |
| LDL cholesterol | 2.9 (1.1) | 2.8 (1.1) | 2.9 (1.1) | -0.32 – 1.31 | 0.41 |
| HDL cholesterol | 1.3 (0.56) | 1.28 (0.58) | 1.37 (0.54) | 0.16 – 0.21 | 0.09 |
| Triglycerides | 1.5 (0.88) | 1.52 (0.89) | 1.50 (0.88) | -0.16 – 0.19 | 0.88 |

\*Significant p-value; †As defined in methodology and Reference [35]; BP, Blood pressure; CI, Confidence interval; DM, Diabetes mellitus. FBG, Fasting blood glucose; GFR, Glomerular filtration rate; HbA1c, Glycated hemoglobin; HDL, High density lipoprotein; LDL, Low density lipoprotein; N/A, Not applicable; SD, Standard deviation; WC, Waist circumference; CKD indicates GFR ≤ 59ml/min/1.73M<sup>2</sup>

### Types of diabetes, duration and treatment

Thirty-Nine (9.7%) participants had type 1 DM and 364 (90.3%) had type 2 DM. Mean age (±SD) at DM diagnosis was 24.2±6.1 years for type 1 DM and 47.11±9.4 years for type 2 DM (95% CI: 19.8–25.9, *P* < 0.001), while median DM duration was 77 and 84 months, respectively. DM care at enrolment involved dietary control alone (1.7%), dietary control with oral GLD only (61.8%), dietary control with insulin only (9.2%), and dietary control combined with oral GLD and either insulin (26.8%) or semaglutide (0.24%).

### Outcome measures

#### 1. Rates and patterns of dyslipidemia

Dyslipidemia was present in 240 (59.6%) participants, comprising 84 males and 156 females (Table 2). The mean age (±SD) of those with dyslipidemia was 53.5±12.8 years, with males being significantly older than females (55.8±11.5 years vs. 52.2±13.3 years, respectively; 95% CI: 0.23–7.0, *P* = 0.036). Lipoprotein levels of dyslipidemia showed a low HDLC in 69.6%, high TC in 26.2%, high LDLC in 25.0%, and high TG in 18.7% participants (Fig 2).

**Fig 2.**
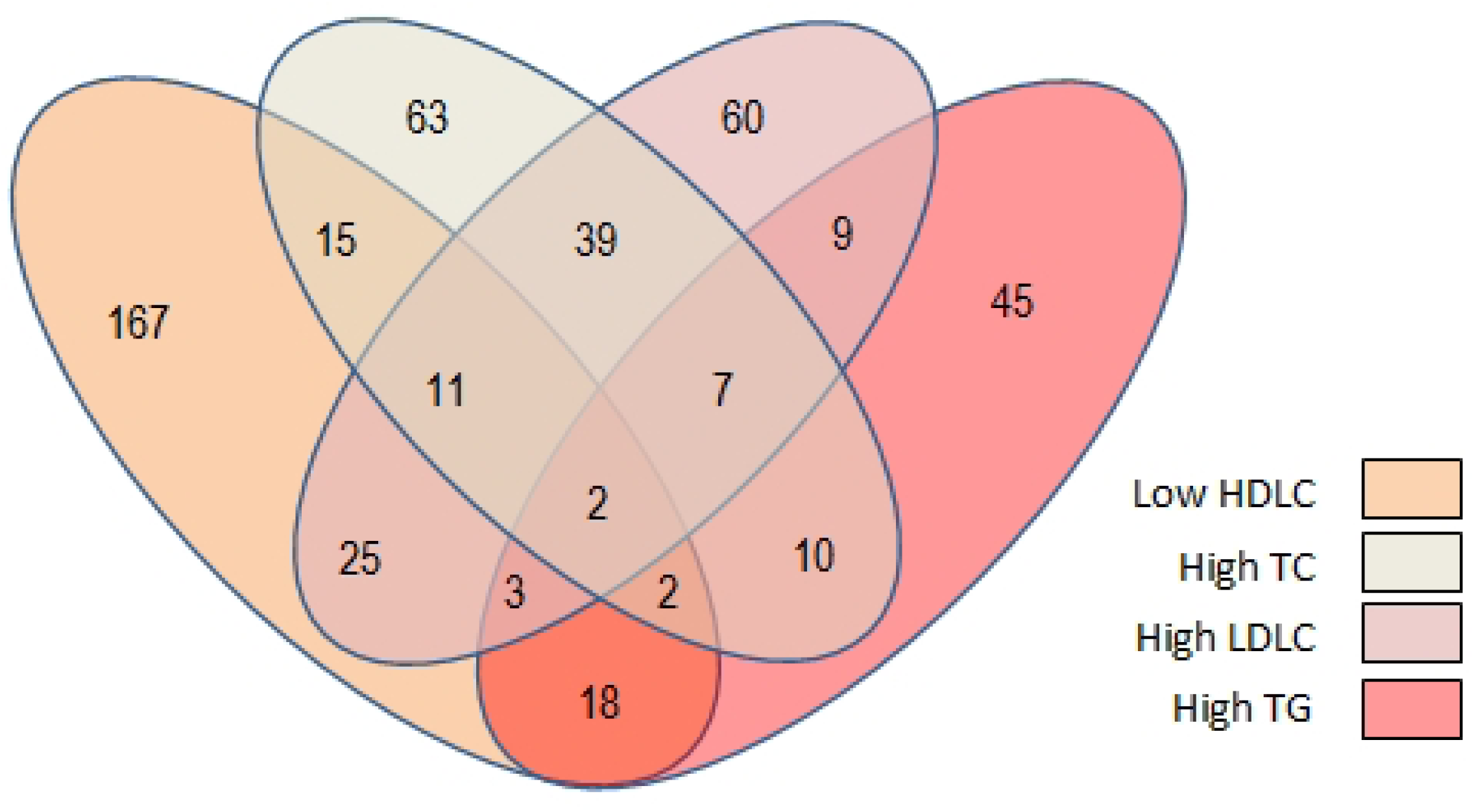
Four-set Venn diagram for lipoprotein levels of dyslipidemia in 240 participants. HDLC, High density lipoprotein cholesterol; LDLC, Low density lipoprotein cholesterol; TC, Total cholesterol; TG, Triglycerides

Using chi square tests, we found that participant characteristics associated with dyslipidemia were the female gender (65% females vs. 35% males; χ^2^ = 9.39, *P* = 0.003), obesity (58.6% vs. 43.7% respectively; χ^2^ = 8.51, *P* = 0.004) and proteinuria (44.8% vs. 32.5% respectively; χ^2^ = 6.074, P < 0.017), but not CKD, age > 50 years, poor glycemic control or other variables (Table 3). Although the rate of dyslipidemia tended to be lower in those with university education, the difference was not significant (43.7% vs. 53.4%; χ^2^ = 3.61, *P* = 0.067). We performed a logistic regression analysis, where dyslipidemia was the dependent variable, and participant characteristics implicated in univariate analysis and in previous studies, were the independent variables. We found that only female gender (OR = 1.74, *P* = 0.022) and proteinuria (OR = 2.26, *P* = 0.004) were independently associated with dyslipidemia after controlling for the effects of confounders.

**Table 3:**
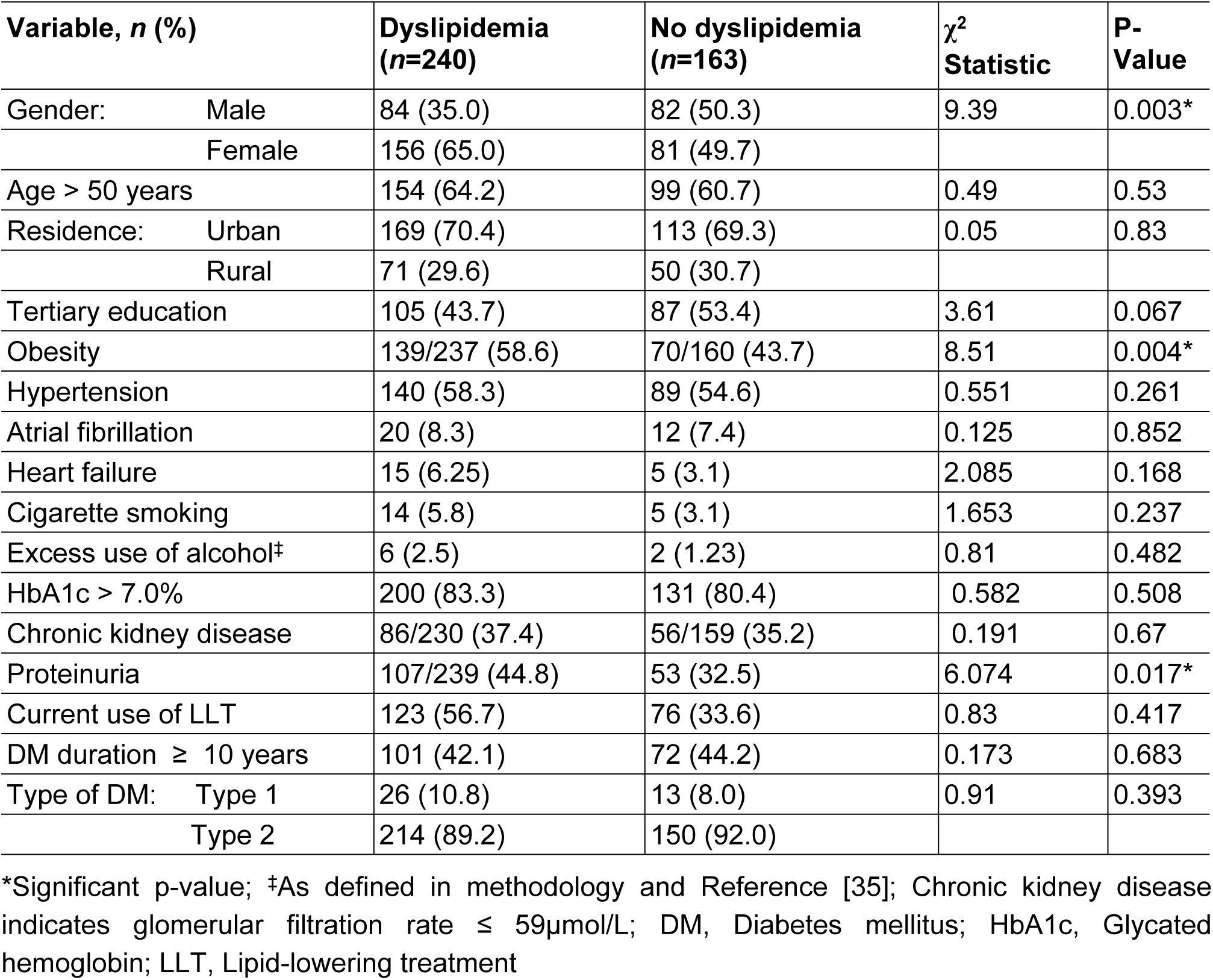
Chi square tests of associations between participant variables and dyslipidemia.

| Variable, *n* (%) | Dyslipidemia<br>(*n*=240) | No dyslipidemia<br>(*n*=163) | $\chi^2$<br>Statistic | P-Value |
| --- | --- | --- | --- | --- |
| Gender: Male | 84 (35.0) | 82 (50.3) | 9.39 | 0.003* |
| Female | 156 (65.0) | 81 (49.7) |  |  |
| Age > 50 years | 154 (64.2) | 99 (60.7) | 0.49 | 0.53 |
| Residence: Urban | 169 (70.4) | 113 (69.3) | 0.05 | 0.83 |
| Rural | 71 (29.6) | 50 (30.7) |  |  |
| Tertiary education | 105 (43.7) | 87 (53.4) | 3.61 | 0.067 |
| Obesity | 139/237 (58.6) | 70/160 (43.7) | 8.51 | 0.004* |
| Hypertension | 140 (58.3) | 89 (54.6) | 0.551 | 0.261 |
| Atrial fibrillation | 20 (8.3) | 12 (7.4) | 0.125 | 0.852 |
| Heart failure | 15 (6.25) | 5 (3.1) | 2.085 | 0.168 |
| Cigarette smoking | 14 (5.8) | 5 (3.1) | 1.653 | 0.237 |
| Excess use of alcohol‡ | 6 (2.5) | 2 (1.23) | 0.81 | 0.482 |
| HbA1c > 7.0% | 200 (83.3) | 131 (80.4) | 0.582 | 0.508 |
| Chronic kidney disease | 86/230 (37.4) | 56/159 (35.2) | 0.191 | 0.67 |
| Proteinuria | 107/239 (44.8) | 53 (32.5) | 6.074 | 0.017* |
| Current use of LLT | 123 (56.7) | 76 (33.6) | 0.83 | 0.417 |
| DM duration ≥ 10 years | 101 (42.1) | 72 (44.2) | 0.173 | 0.683 |
| Type of DM: Type 1 | 26 (10.8) | 13 (8.0) | 0.91 | 0.393 |
| Type 2 | 214 (89.2) | 150 (92.0) |  |  |

\*Significant p-value; ‡As defined in methodology and Reference [35]; Chronic kidney disease indicates glomerular filtration rate $\leq 59\mu\text{mol/L}$ ; DM, Diabetes mellitus; HbA1c, Glycated hemoglobin; LLT, Lipid-lowering treatment

#### 2. Proportion of patients treated for dyslipidemia

At enrolment, 202 participants (50.1%) were taking LLTs, of whom 123 had dyslipidemia. Thus, LLT use for dyslipidemia was 123/240 (51.3%), which included statins only (49.6%), clofibrate only (1.7%), and a statin combined with clofibrate (1.2%). The 79 participants without dyslipidemia took only statins, and none with dyslipidemia took niacin, ezetimibe, bempedoic acid, inclisiran, PCSK9 inhibitors or icosapent ethyl. Chi square tests showed no differences in the number of participants with dyslipidemia who did, and did not use LLTs (51.2% vs. 45.6%, respectively; χ^2^ = 0.83, *P* = 0.417).

#### 3. Proportion of patients that attained LDLC goal and target

Among 403 participants, 69 (17.1%) had attained the LDLC goal and target for the primary prevention of CVD in people with DM. Chi square tests of association showed no significant differences in LDLC targets with respect to gender (18.9% males vs. 15.2% females; χ^2^ = 1.513, *P* = 0.23), university education (16.7% educated vs. 17.5% uneducated; χ^2^ = 0.053, *P* = 0.89), glycemic control (15.3% good vs. 17.5% poor; χ^2^ = 0.21, *P* = 0.73), use of LLT (18.1% vs. 16.2%; χ^2^ = 0.26, *P* = 0.69) or other participant variables.

## Discussion

The study found a mean participant age of 53.2 years, with females being younger than males. The overall prevalence of dyslipidemia was 59.6%, with the rate significantly higher in females than males, in the obese than the non-obese, and in those with proteinuria. The prevalence rate in this study was comparable to earlier findings in Nigeria, but higher than the 43.5% rate among type 1 DM patients in Spain, and lower than the 62.3% rate among type 1 and type 2 DM patients in China [16,27,38]. Consistent with our findings of a higher female prevalence of both obesity and dyslipidemia, the U.S. National Health and Nutrition Examination Survey (NHANES) also found women to be more obese than men, at 40% and 35% rates, respectively [39]. Obesity being a cause of dyslipidemia, the higher rate of dyslipidemia in our female participants was likely due to the higher rate of obesity.

In this study, dyslipidemia was also associated with proteinuria, but not with CKD, poor glycemic control, DM duration more than 10 years, university education, excess use of alcohol, or other demographic or clinical characteristics of participants. Previous studies have associated dyslipidemia with diabetic kidney disease and higher risks of CVD [40,41]. In the study by Hirano et al, diabetic kidney disease, with and without proteinuria, both correlated with higher risks of CVD and higher levels of serum lipoproteins, while proteinuria was independently associated with even higher risks of these outcomes [40]. We found dyslipidemia was similarly associated with proteinuria, but not with CKD assessed by renal function. This disparity could be due to a different methodology than the Japanese study. Whereas we used only the GFR as a measure of renal function, Hirano et al. assessed both GFR and the urinary albumin-to-creatinine ratio that is reportedly more sensitive to kidney injury. Moreover, Nigeria has the fourth highest prevalence of CKD worldwide, after Iran, Panama and Malaysia [42]. With uncontrolled hypertension, post-streptococcal glomerulonephritis, renal tuberculosis and other infections contributing much to the CKD burden in Nigeria, a large proportion of our patients with CKD may not have had diabetic kidney disease [43].

The most common form of dyslipidemia in this study was a low HDLC, and the least common was a raised TG. Previous studies from Nigeria have yielded mixed results. While some studies have reported low HDLC to be most common in the Southeast and Southwest regions, studies from the Northwest and North-Central regions have reported high LDLC and high TG to be more common [19,27,28]. Different dietary practices and genetic factors influencing lipid absorption, transport and metabolism among the various ethnic groups of Nigeria could explain these disparities. For instance, consumption of palm oil, a common component of the southern diet, is associated with higher levels of LDLC compared to diets prepared with vegetable oils low in saturated fat and more widely consumed in northern Nigeria [44]. On the other hand, regular consumption of cheese, which is prevalent in the northern regions, is known to lower LDLC but also HDLC when compared to regular consumption of butter [45], which is common in the south.

We observed that 51.3% participants with dyslipidemia were taking statins and other LLTs, which was similar to the 53.8% treatment rate in Arsi, Ethiopia, but lower than the 68% rate in Northwest China [26,46]. In contrast, studies from Turkey and Mexico reported lower treatment rates of 42.4% and 23.3%, respectively [47,48]. Few studies from Nigeria have reported treatment rates for dyslipidemia in DM patients. A multicenter study has found only 12.6% patients were on statin treatment for dyslipidemia, although the rate of dyslipidemia was not stated [49]. Another multinational study of lipid management (the INTERASPIRE study) has found a treatment rate of 68.6% among Nigerians, of whom 40% were DM patients [25]. In that study, LLT use in Nigeria included statins (67.3%), fibrates (0.4%), and a statin-ezetimibe combination (1.3%). Although we found a lower treatment rate, the LLTs were similar, and neither study has recorded the use of niacin, bempedoic acid, inclisiran, or PCSK9 inhibitors such as alicorumab. Economic factors could explain the non-use of these LLTs despite their proven benefits in those who failed intensive statin therapy or are intolerant to the side effects [15]. With health insurance limited in Nigeria, most patients with dyslipidemia tend to use out-of-pocket funds to pay for prescription medicines, including LLTs. Thus, most patients may afford cheaper generic forms of statins, but not newer LLTs lacking off-patent substitutes.

We found no association between LLT use and dyslipidemia, which was expected. Beside their use in dyslipidemia, statins are also recommended for secondary prevention of CVD in people with DM, as well as primary prevention in those at very high risks [15]. Our study protocol also restricted dyslipidemia to abnormal serum lipids, regardless of current use of LLT for a previous diagnosis of dyslipidemia. While this has excluded participants who self-medicated with LLTs on false assumptions of dyslipidemia, it did not exclude wrong use of LLT, which could cause an effect bias when comparing LLT use in those with, and those without, dyslipidemia.

Finally, we found that only 17.1% of all participants had attained the LDLC goal and target recommended by the European Society of Cardiology for primary prevention of CVD in people with DM [15]. Although some participants needed secondary prevention due to previous CVD, they were too few for separate analysis, and were jointly assessed for primary prevention. Our findings were similar to Uloko et al, who found only 21.1% males and 13.9% females meeting the HDLC targets of the American Diabetes Association [49]. In contrast, a Chinese study reported a higher rate of 43.1% DM patients meeting LDLC targets [45]. Li et al. also reported that lower levels of HbA1c and current use of LLT each correlated with attainment of LDLC target. On the contrary, we did not find these correlations. This could arise for several reasons, such as poor patient adherence to treatment regimens, failure of clinicians to test for dyslipidemia, or failure to prescribe the correct form and dosage of LLTs recommended in clinical practice guidelines.

### Limitations of the study

Some participants had poor recollections of a diagnosis of PTE, which could under-estimate the rate of previous CVD. However, this didn’t impact on the main outcome measures. Secondly, we restricted the definition of dyslipidemia to abnormal levels of serum lipoproteins tested during the study, regardless of previous history of dyslipidemia. While this has excluded false dyslipidemia based on participant self-diagnosis, it may have raised the proportion of those lacking dyslipidemia but taking LLTs at enrolment. We mitigated that by emphasizing on dyslipidemia diagnosed by physicians based on lipid profile tests, and use of LLT prescribed by physicians only. Lastly, a few participants had missing data on BMI, waist circumference and renal function tests. We addressed that by excluding missing counts from denominators during statistical analysis.

### Summary and conclusion

In this study of DM patients in northern Nigeria, we found a 59.6% prevalence of dyslipidemia that was similar to earlier findings in Nigeria and elsewhere. Participant characteristics independently associated with dyslipidemia were female gender and proteinuria. Although half of those with dyslipidemia took LLTs, this did not differ from those lacking dyslipidemia. LLTs comprised of statins and clofibrate, but not niacin, ezetimibe, bempedoic acid, inclisiran or PCSK9 inhibitors, probably due to higher costs of branded medicines. All participants were DM clinic patients, but only 17.1% attained the LDLC target recommended for primary prevention of CVD. In conclusion, dyslipidemia is prevalent among DM patients in northern Nigeria, and is mostly untreated. There is a need for better physician adherence to clinical practice guidelines and improved patient access to dyslipidemia treatment, especially in females, in northern Nigeria.

## Author Contributions

**Conceptualization:** Nura H. Alkali, Andrew E. Uloko, Mohammed R. Bello, Maria A. Garba, Ayuba Mugana, Ijuptil Chiroma

**Data curation:** Nura H. Alkali, Andrew E. Uloko, Maria A. Garba, Zira G. Vandi, Muhammad A. Saad, Ayuba Mugana, Ijuptil Chiroma, Umar F. Abdullahi, Lumsami Shadrach, Usman A. Nuhu

**Formal analysis:** Nura H. Alkali

**Investigation:** Nura H. Alkali, Andrew E. Uloko, Adamu G. Bakari, Mohammed R. Bello, Maria A. Garba, Muhammad A. Saad, Zira G. Vandi, Ayuba Mugana, Ijuptil Chiroma, Umar F. Abdullahi, Ibrahim I. Haladu, Lumsami Shadrach, Usman A. Nuhu, Godiya I. Dare

**Methodology:** Nura H. Alkali, Abubakar S. Muhammad

**Project administration:** Nura H. Alkali, Andrew E. Uloko, Godwin O. Osaigbovo, Godiya I. Dare

**Supervision:** Mohammed R. Bello, Abubakar S. Muhammad

**Validation:** Andrew E. Uloko, Godwin O. Osaigbovo, Adamu G. Bakari, Maria A. Garba, Garba M. Fika

**Visualization:** Nura H. Alkali

**Writing – original draft:** Nura H. Alkali

**Writing – review & editing:** Nura H. Alkali, Andrew E. Uloko, Godwin O. Osaigbovo, Adamu G. Bakari, Mohammed R. Bello, Maria A. Garba, Garba M. Fika, Abubakar S. Muhammad, Muhammad A. Saad, Zira G. Vandi, Umar F. Abdullahi, Ayuba Mugana, Ijuptil Chiroma, Ibrahim A. Haladu, Lumsami Shadrach, Usman A. Nuhu, Godiya I. Dare

## Data availability statement

Data are available from the corresponding author upon reasonable request.

## Acknowledgments

We appreciate the Nurse Managers of DM Clinics at the study sites and resident doctors who assisted in data collection, especially Matron Frama Ali at ATBUTH, Matron Hajara Mbaya at UMTH, Dr. Kabiru Audi at ATBUTH and Dr. Raphael Faruna at MAUTH.

